# Non-invasive ²³Na MRI Identifies Functionally Distinct Cyst Phenotypes in Autosomal Dominant Polycystic Kidney Disease

**DOI:** 10.64898/2026.07.30.26359242

**Authors:** Judith Schirmer, Laurent Ruck, Anke Dahlmann, Peter Linz, Katharina Tkotz, Jordan M. Höhn, Raluca Ursu, Andre Kraus, Silke Haerteis, Barbara Nübel, Marc Saake, Bernd Wullich, Mario Schiffer, Michael Uder, Björn Buchholz, Armin M. Nagel, Christoph Kopp

## Abstract

Autosomal dominant polycystic kidney disease (ADPKD) is characterized by cysts from different nephron segments, yet their origin remains inaccessible to in vivo imaging. Building on ex vivo studies that cyst sodium concentration ([Na⁺]) may reflect tubular origin, we developed a non-invasive ^23^Na magnetic resonance imaging (MRI) approach at 7 Tesla to phenotype ADPKD cysts in vivo based on cyst sodium. In nephrectomized ADPKD kidneys, cyst fluid [Na^+^] closely matched ^23^Na MRI signal intensities, revealing two distinct cyst phenotypes: serum-like high-[Na^+^] cysts and low-[Na^+^] cysts. High-[Na^+^] cysts expressed the proximal tubular marker sodium-glucose cotransporter-2, whereas low-[Na^+^] cysts expressed markers of distal tubules and collecting ducts. The vasopressin V2 receptor (V2R), the therapeutic target of tolvaptan, was detected exclusively in low-[Na^+^] cysts, linking sodium phenotype to therapeutic target expression. In vivo, ²³Na MRI enabled classification of 2,299 cysts in 20 ADPKD patients. High-[Na^+^] cysts were 1.8-fold more frequent overall. Anatomical cyst localization did not predict sodium phenotype, underscoring the need for functional imaging rather than structural inference. Patients exhibited marked interindividual variation in cyst composition, with low-[Na^+^] cysts comprising 4.6% to 83.9% of all cysts. Given the exclusive expression of V2R in low-[Na^+^] cysts, this heterogeneity may influence disease progression and therapeutic responsiveness. These findings establish ²³Na MRI as a non-invasive method to phenotype ADPKD cysts in vivo and provide a functional imaging approach with potential relevance for stratifying patients and guiding future therapeutic interventions.

## INTRODUCTION

Autosomal dominant polycystic kidney disease (ADPKD) is the most common form of inherited cystic disease in humans and accounts for 5 – 10% of end-stage kidney disease cases (*1,2*). In ADPKD, renal cysts can originate from all segments of the nephron and collecting duct (*3*). Consequently, cysts in ADPKD do not form a homogeneous population but express a wide variety of tubule markers, and their cyst fluid shows a heterogeneous composition (*4,5*). Ex vivo data suggest a highly variable sodium concentration ([Na^+^]) in renal ADPKD cysts (*6–8*). However, data on cyst fluid composition in relation to tubule markers are lacking. Furthermore, earlier studies have indicated that cyst [Na^+^] reflects its tubular origin (*6*). Electrophysiological studies have further suggested that low-and high-[Na^+^] cysts differ in epithelial transport properties consistent with distal and proximal nephron origin, respectively (*9*). Despite this knowledge, no systematic in vivo data on the incidence, abundance and distribution of cysts with different [Na^+^] exists. This gap reflects the lack of a clinically applicable technique to assess cyst [Na^+^] non-invasively in humans. The distinction between the different cyst types may have direct therapeutic implications. Tolvaptan, currently the only approved medical therapy for slowing cyst growth in ADPKD patients (*10*), acts through the vasopressin V2 receptor (V2R), which is localized to the distal nephron and collecting duct. Consequently, only cysts originating from these nephron segments are expected to respond directly to V2R inhibition.

We therefore investigated whether cyst sodium concentration defines biologically distinct cyst populations, reflects tubular origin, and can be assessed non-invasively using in vivo ^23^Na MRI to enable imaging-based classification of ADPKD cysts.

## RESULTS

### Cyst [Na^+^] can be reliably assessed by ^23^Na MRI in ADPKD kidneys ex vivo

Initially, eight polycystic kidneys from ADPKD patients – seven on haemodialysis and one kidney transplant recipient – were assessed ex vivo immediately after nephrectomy by 7 Tesla ^23^Na MRI. Representative anatomical (^1^H) and sodium (^23^Na) images of an ADPKD kidney are depicted in **Fig. 1A**. ^23^Na MRI could clearly identify areas of low-and high-[Na^+^] within the kidneys that could be ascribed to kidney cysts in the ^1^H images. Cysts were categorized according to the ^23^Na MRI-detected [Na^+^] and corresponding cyst fluids were assessed after aspiration. Results from laboratory assessment of cyst fluid were either comparable to serum [Na^+^] (141 mmol/L, IQR 138-144, n = 39) or very low (16 mmol/L, IQR 10-36, n = 32, p<0.001). A cutoff [Na^+^] of 100 mmol/L was used to discriminate between the two cyst types as proposed by other researchers (*7,11*). **Table 1** summarizes the characteristic composition of the acquired cyst fluid samples from low-[Na^+^] and high-[Na^+^] cysts (**Fig. 1A**). ^23^Na MRI was able to reliably distinguish between the two cyst entities (low-[Na^+^]: 31 mmol/L, IQR 24-36, n = 32 vs. high-[Na^+^]: 128 mmol/L, IQR 118-147, n = 37, p < 0.001, **Fig. 1B**). Only one cyst showed a discrepancy between MRI and laboratory result whereas two cysts could not be assigned during postprocessing of the images. A close correlation was found between both measurements - ^23^Na MRI and direct assessment of [Na^+^] by ion selective electrode (r = 0.76, p < 0.001, n = 69, Spearman’s rank correlation). Both measurements yielded [Na^+^] in a similar range (^23^Na MRI vs. cyst fluid; low-[Na^+^]: 31 mmol/L, IQR 24-36 vs. 16 mmol, IQR 10-36, n = 32; p = 0.09 and high-[Na^+^]: 128 mmol/L, IQR 118-147 vs. 142 mmol/L, IQR 138-144, n = 37, p = 0.02, **Fig. 1B**). In high-[Na^+^] cysts, [Na^+^] measured by ^23^Na MRI was slightly lower compared to the corresponding laboratory data; however, this discrepancy did not affect the ability of ^23^Na MRI to discriminate between both cyst types (**Fig. 1B**).

**Fig. 1:**
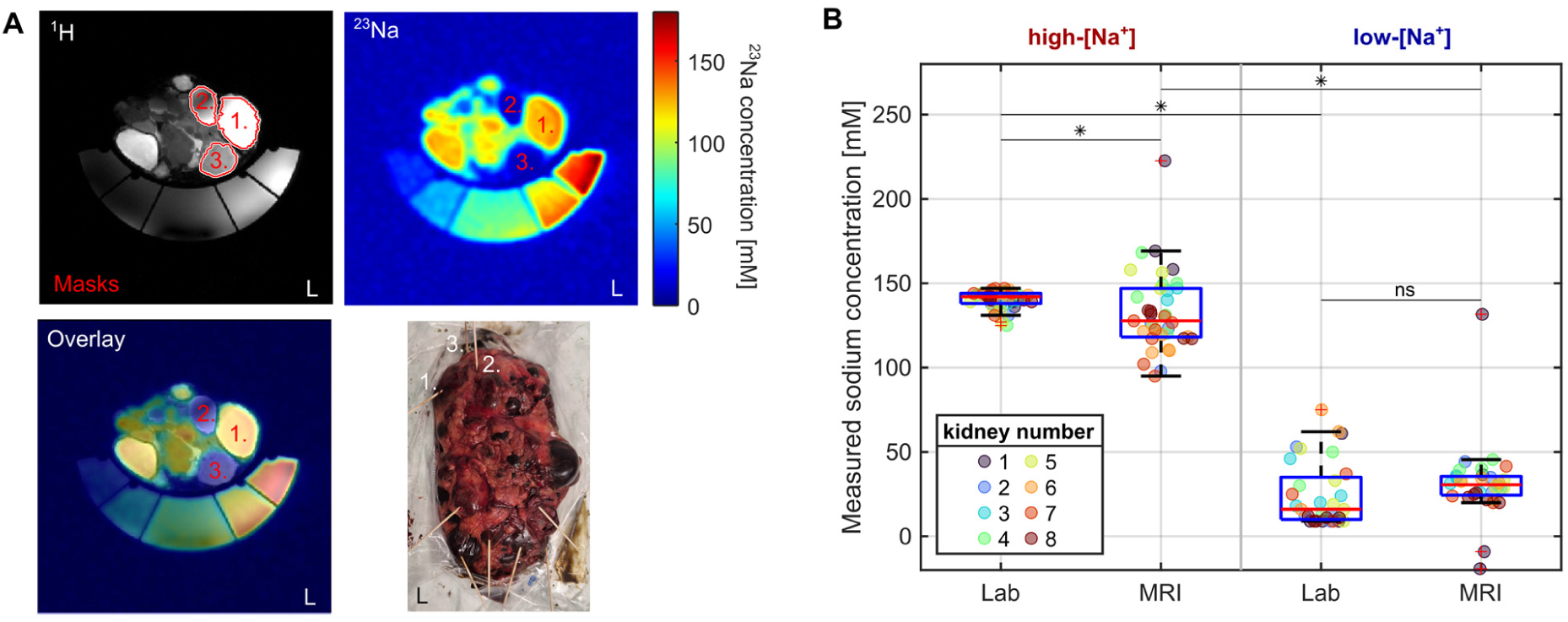
Assessment of cyst [Na^+^] by ^23^Na MRI ex vivo and laboratory validation. **A** Representative anatomical ^1^H and ^23^Na magnetic resonance (MR) images of a nephrectomized kidney acquired at 7 Tesla (axial plane). References with 30, 60, 90, 120 and 150 mmol/L Na^+^ for signal calibration are placed below the kidney. Toothpicks on the ADPKD kidney indicate dedicated punctured cysts. Three example cysts are marked within each image (1.-3.). **B** Comparison of [Na^+^] assessment between ^23^Na MRI and laboratory analysis of punctured cysts from eight nephrectomized kidneys. 69 cysts were evaluated by both methods, 32 with high and 37 with low-[Na^+^] as defined by laboratory measurements (threshold at 100 mmol/L). Both measurements yielded [Na^+^] in similar range (^23^Na MRI vs. cyst fluid; low-[Na^+^]: 31 mmol/L, vs. 16 mmol, IQR 10-36, n = 32, p = 0.09 and high-[Na^+^]: 128 mmol/L, vs. 142 mmol/L, n = 37, p = 0.02, Wilcoxon signed-rank test). ^23^Na MRI could clearly discriminate between both cyst types (low-[Na^+^]: 31 mmol/L, n = 32 vs. high-[Na^+^]: 128 mmol/L, n = 37, p < 0.001, Mann-Whitney U test). Only one cyst showed a mismatch between laboratory and ^23^Na MRI analysis. * p < 0.05; ns, not significant.

**Table 1:**
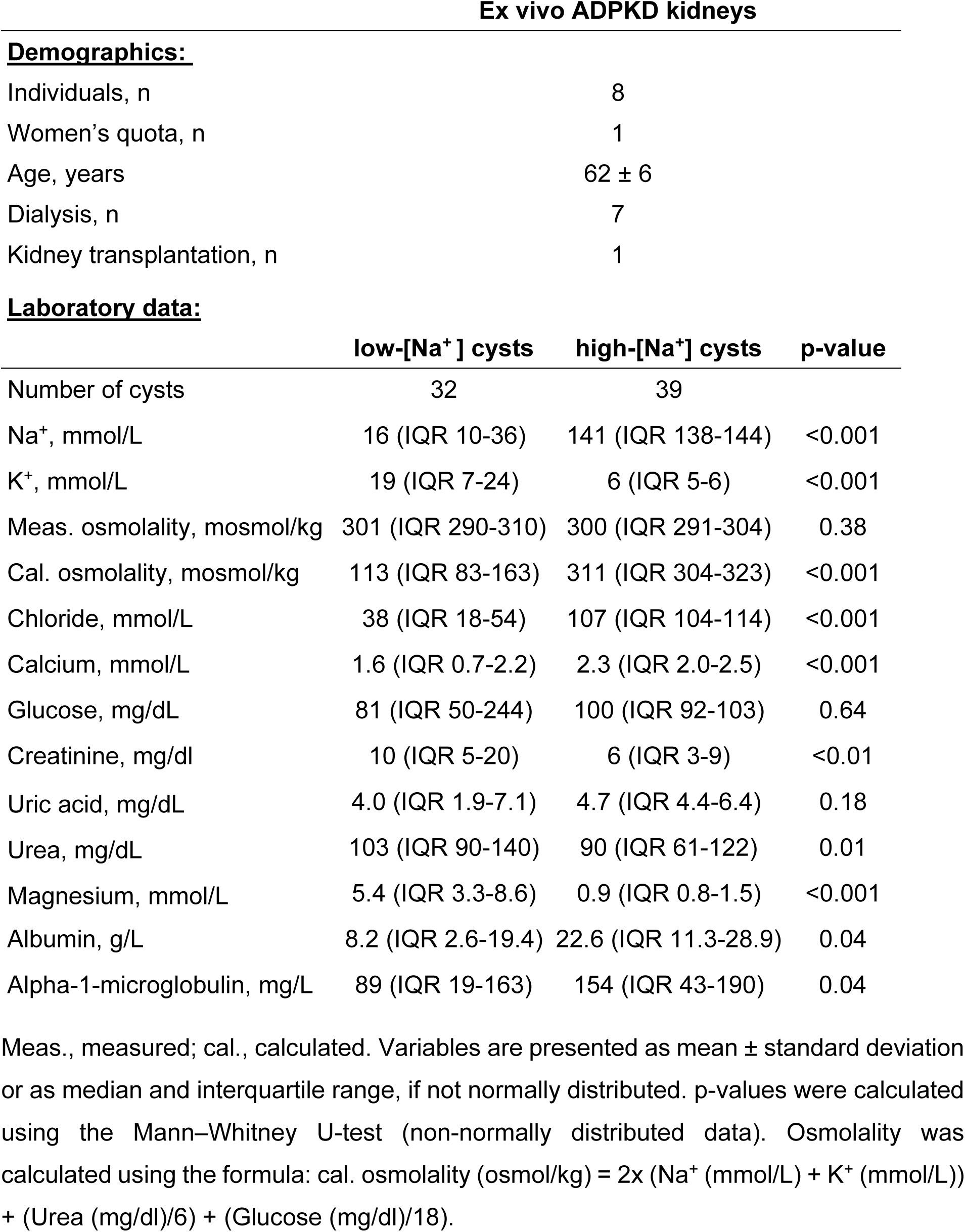
Demographics of nephrectomized patients and laboratory data of cyst fluid.

| Ex vivo ADPKD kidneys |  |  |  |
| --- | --- | --- | --- |
| **<u>Demographics:</u>** |  |  |  |
| Individuals, n |  | 8 |  |
| Women's quota, n |  | 1 |  |
| Age, years |  | 62 ± 6 |  |
| Dialysis, n |  | 7 |  |
| Kidney transplantation, n |  | 1 |  |
| **<u>Laboratory data:</u>** |  |  |  |
|  | low-[Na<sup>+</sup>] cysts | high-[Na<sup>+</sup>] cysts | p-value |
| Number of cysts | 32 | 39 |  |
| Na<sup>+</sup>, mmol/L | 16 (IQR 10-36) | 141 (IQR 138-144) | <0.001 |
| K<sup>+</sup>, mmol/L | 19 (IQR 7-24) | 6 (IQR 5-6) | <0.001 |
| Meas. osmolality, mosmol/kg | 301 (IQR 290-310) | 300 (IQR 291-304) | 0.38 |
| Cal. osmolality, mosmol/kg | 113 (IQR 83-163) | 311 (IQR 304-323) | <0.001 |
| Chloride, mmol/L | 38 (IQR 18-54) | 107 (IQR 104-114) | <0.001 |
| Calcium, mmol/L | 1.6 (IQR 0.7-2.2) | 2.3 (IQR 2.0-2.5) | <0.001 |
| Glucose, mg/dL | 81 (IQR 50-244) | 100 (IQR 92-103) | 0.64 |
| Creatinine, mg/dl | 10 (IQR 5-20) | 6 (IQR 3-9) | <0.01 |
| Uric acid, mg/dL | 4.0 (IQR 1.9-7.1) | 4.7 (IQR 4.4-6.4) | 0.18 |
| Urea, mg/dL | 103 (IQR 90-140) | 90 (IQR 61-122) | 0.01 |
| Magnesium, mmol/L | 5.4 (IQR 3.3-8.6) | 0.9 (IQR 0.8-1.5) | <0.001 |
| Albumin, g/L | 8.2 (IQR 2.6-19.4) | 22.6 (IQR 11.3-28.9) | 0.04 |
| Alpha-1-microglobulin, mg/L | 89 (IQR 19-163) | 154 (IQR 43-190) | 0.04 |

Meas., measured; cal., calculated. Variables are presented as mean ± standard deviation or as median and interquartile range, if not normally distributed. p-values were calculated using the Mann–Whitney U-test (non-normally distributed data). Osmolality was calculated using the formula: cal. osmolality (osmol/kg) = 2x (Na<sup>+</sup> (mmol/L) + K<sup>+</sup> (mmol/L)) + (Urea (mg/dl)/6) + (Glucose (mg/dl)/18).

### Cyst [Na^+^] reflects tubular origin in ADPKD

A total of 41 renal cysts from six of the eight ADPKD nephrectomy specimens were additionally analyzed for tubular segment identity. Based on intraluminal Na^+^ measurements, 22 cysts were classified as high-[Na^+^] and 19 as low-[Na^+^] cysts.

Low-[Na^+^] cysts displayed a highly consistent distal/collecting-duct phenotype. Aquaporin-2 (AQP2) was positive in 17 of 19 cysts (89.5%), and V2R in 13 of 19 cysts (68.4%), indicating robust collecting-duct differentiation. Sodium-chloride cotransporter (NCC) was detected in 6 of 19 cysts (31.6%). Notably, NCC positivity never occurred in isolation but always in combination with AQP2 and/or V2R. Three cysts showed an NCC+/AQP2+/V2R+ profile, consistent with a connecting tubule (CNT)-to-early collecting duct (CD) identity, whereas the remaining three cysts displayed an NCC+/AQP2+/V2R– pattern, compatible with late distal convoluted tubule (DCT2)/CNT differentiation. None of the low-[Na^+^] cysts expressed sodium-glucose cotransporter-2 (SGLT2).

In contrast, high-[Na^+^] cysts uniformly lacked distal markers; none of the 22 high-[Na^+^] cysts expressed AQP2, V2R, or NCC. Instead, SGLT2 was positive in 13 of 22 high-[Na^+^] cysts (59.1%), supporting a proximal-tubule origin in a substantial subset. The remaining high-[Na^+^] cysts were negative for all segment-specific markers. Additional proximal markers (AQP1, Megalin, Lotus lectin) did not provide further discriminatory value. AQP1 was positive in only three cysts and overlapped almost entirely with SGLT2; Megalin showed aberrant nuclear staining, and Lotus lectin produced diffuse unspecific labelling (**Fig. 2 and Supplementary Fig. 1**).

**Fig. 2:**
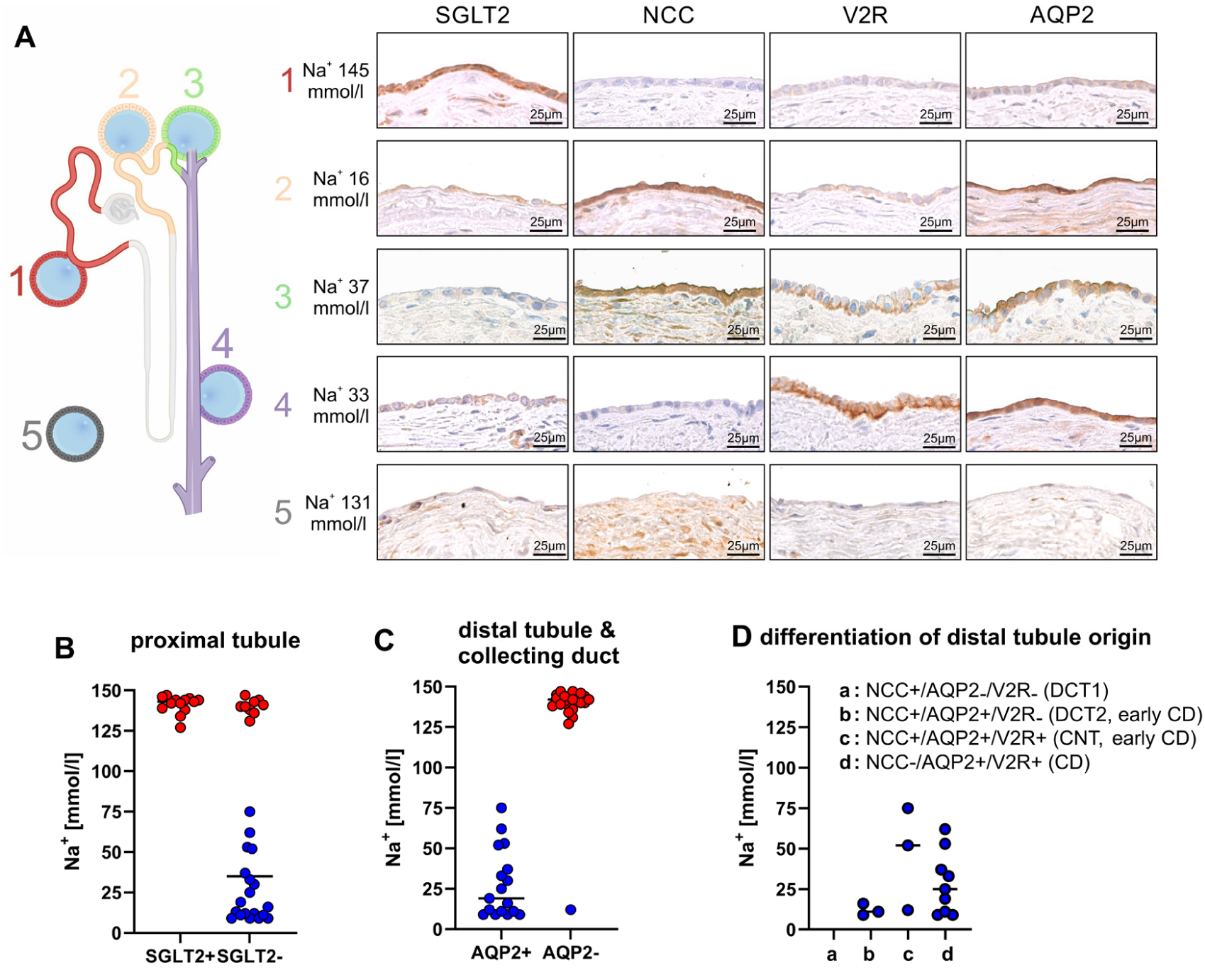
Low-and high-[Na^+^] cysts can be assigned to specific tubular segments. Tubular markers of cysts excised following ^23^Na MRI assessment. **A** Representative stainings of low-[Na^+^] and high-[Na^+^] cysts. High-[Na^+^] cysts either stained positively for the proximal tubular marker sodium-glucose cotransporter-2 (SGLT2) (59.1% of cases, No. 1), or lacked expression of any tubular markers (No 5). Low-[Na^+^] cysts expressed aquaporin-2 (AQP2) in 89.5% and vasopressin V2 receptor (V2R) in 68.4% of cases, combined with sodium-chloride cotransporter (NCC) in 31.6% of cysts (No 2,3,4). **B** None of the low-[Na^+^] cysts (blue) showed a positive staining for the proximal tubular marker SGLT2. **C** Distal tubular markers could not be detected in high-[Na^+^] cysts (red). **D** Based on the combined expression of AQP2, V2R and NCC low-[Na^+^] cysts were assigned to the late distal convoluted tubule (DCT), connecting tubule (CNT) and collecting duct (CD).

Together, these data demonstrate a clear dichotomy in cyst identity: low-[Na^+^] cysts exhibit a robust distal/CNT/collecting-duct phenotype, whereas high-[Na^+^] cysts are non-distal and frequently stained positive for the proximal marker SGLT2, consistent with a proximal-like profile. This molecular separation aligns closely with the functional [Na^+^] of the cyst fluid.

### Frequency and interindividual distribution of both cyst [Na^+^] types in vivo

After establishing the methodology and verifying the imaging results, an observational study was conducted in 20 ADPKD patients to categorize the cyst [Na^+^] in vivo. Given the limited spatial resolution of ^23^Na MRI, only cysts with a volume > 1 ml were assessed. Clinical characteristics of the patients are outlined in **Table 2**. In all ADPKD patients, cysts with both high-[Na^+^] and low-[Na^+^] were present. Among the classified cysts, high-[Na⁺] cysts were 1.8-fold more frequent than low-[Na⁺] cysts (n = 1467 vs. n = 832). However, the individual relative abundance of the two different cyst types was highly variable ranging from 4.6% to 83.9% cysts with low-[Na^+^] (**Supplementary Table 3**). **Fig. 3A+B** shows representative images of a patient with mainly high-[Na^+^] cysts (91.6%, Total Kidney Volume (TKV) = 6.58 L/ height-adjusted TKV (htTKV) = 3978 mL/m) compared to a patient with mainly low-[Na^+^] cyst (68.5%, TKV = 5.58 L/ htTKV = 3101 mL/m) in **Fig. 3C**. In this cohort of ADPKD patients, no correlation between the relative amount of low-[Na^+^] cysts and htTKV was found (**Fig. 3D**). Additionally, no association could be detected between the estimated glomerular filtration rate (eGFR) and the percentage of low-[Na^+^] cysts (**Fig. 3E**). However, higher age was significantly associated with an increased percentage of cysts categorized as high-[Na^+^] by ^23^Na MRI (r = 0.57, p < 0.009) (**Fig. 3E**).

**Fig. 3:**
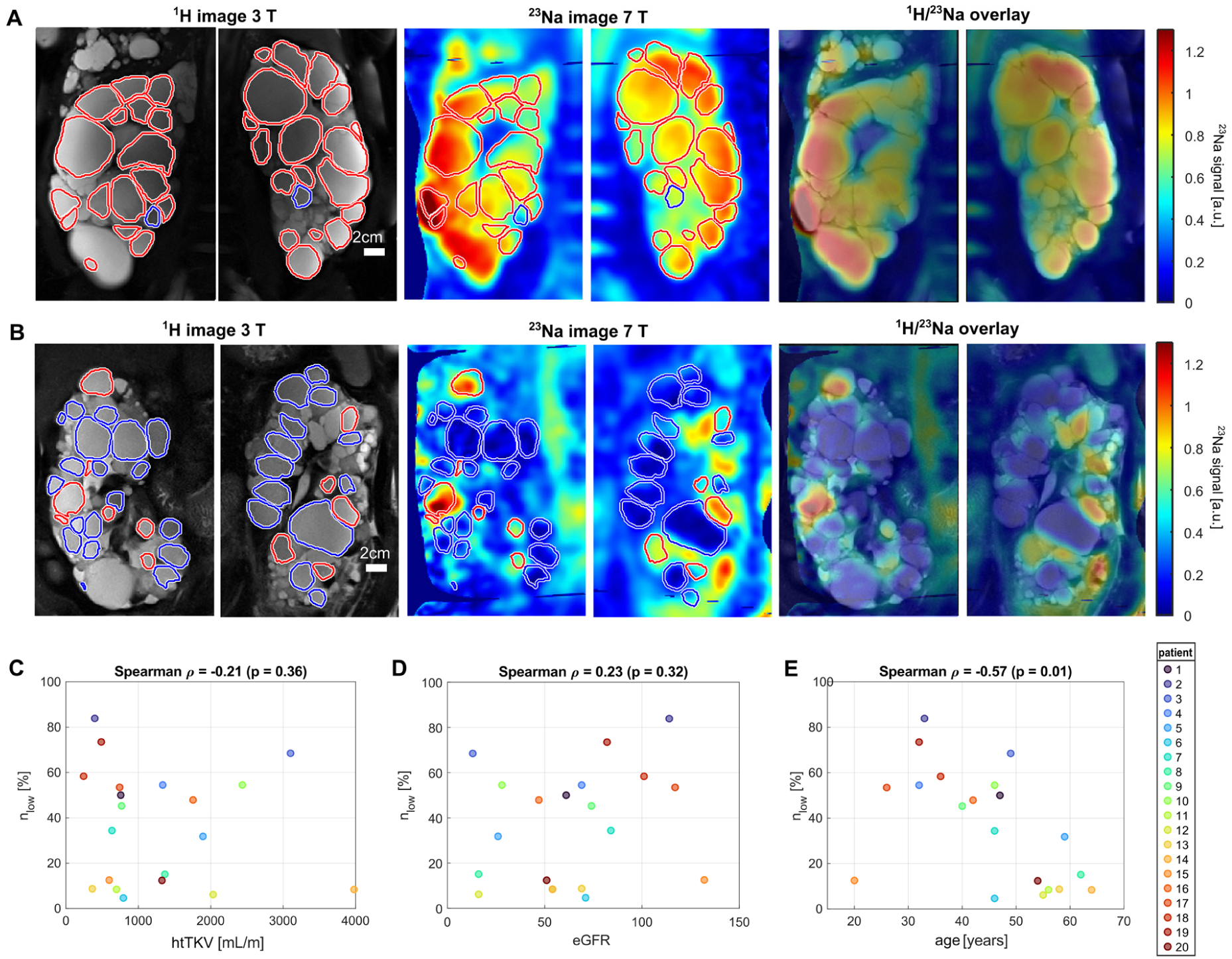
Characterization of cyst [Na^+^] in ADPKD patients in vivo. A/B. Images from two exemplary patients with a distinct percentage of low-and high-[Na^+^] cysts. anatomical ^1^H images acquired at 3 Tesla are shown; alongside ^23^Na MR images obtained at 7 Tesla are depicted. ^23^Na MR images were normalized to the mean signal of the high-[Na^+^] cysts and co-registered to ^1^H MR images. Cyst masks indicate those included in the evaluation. Cyst masks of low-[Na^+^] cysts are shown in blue, high-[Na^+^] cysts in red. Additionally an overlay of ^1^H and ^23^Na MR images is depicted. (**A)** Patient 11 was characterized by a low proportion of low-[Na^+^] cysts (8.4%), whereas 68.5% of the cysts analyzed in patient 3 were classified as low-[Na^+^] (**B)**. **C-E** Marked heterogeneity in the abundance of low-and high-[Na^+^] cysts. Correlation scatter plots between the percentage of low-[Na^+^] cysts (n_low_, [%]) and **(C)** height-adjusted total kidney volume (htTKV), **(D)** estimated glomerular filtration rate (eGFR), and **(E)** patient age are depicted. A significant negative correlation was observed between n_low_ and age (ρ = −0.57, p < 0.009, Spearman), whereas no significant correlations were found with htTKV or annual growth rate. Each dot represents an individual patient.

**Table 2:**
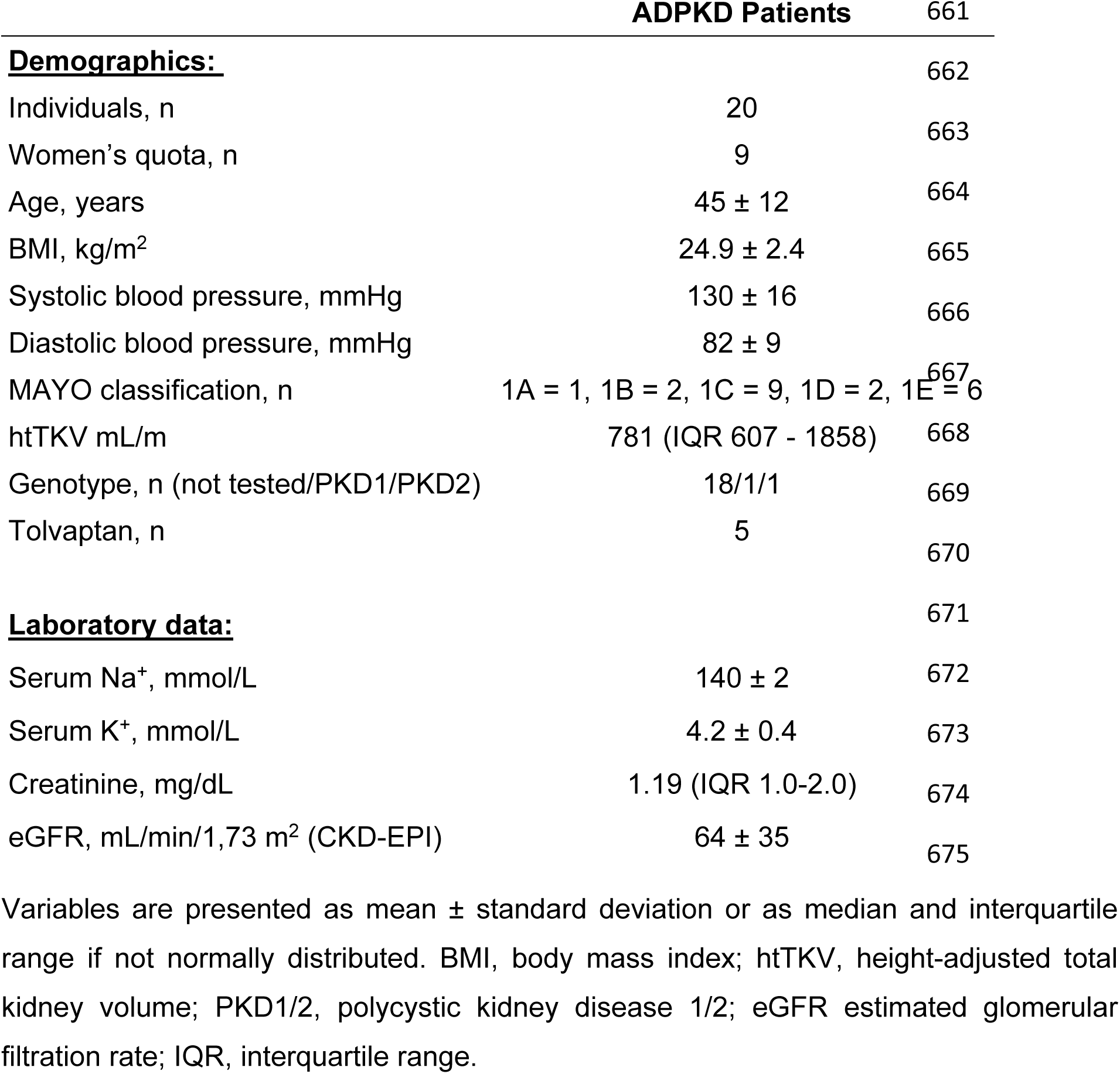
Patient characteristics and laboratory data.

### In vivo size and localization of low-and high-[Na^+^] cysts

To assess whether the cyst size might predict its [Na^+^], we compared the volume of the two different cyst entities (for cysts > 1 mL). The median size of low-[Na^+^] cysts compared to the median size of high-[Na^+^] cysts were significantly different (low-[Na^+^] 2.8 mL vs. high-[Na^+^] 3.3 mL, p < 0.01) and above a volume of 72 mL solely high-[Na^+^] cysts were present in our cohort (**Fig. 4A**).

**Fig. 4:**
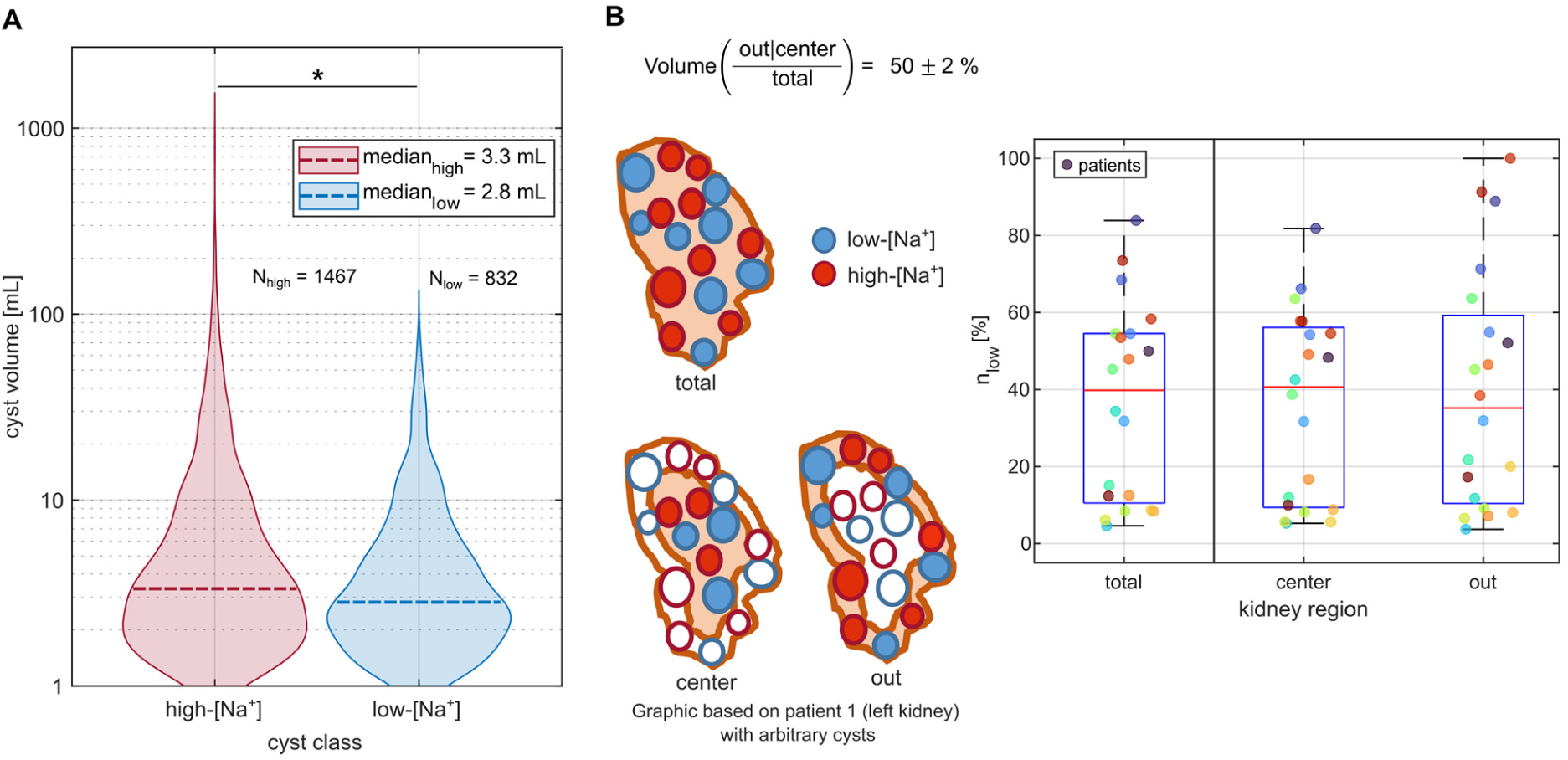
Volume and spatial distribution of low-and high-[Na^+^] cysts. **A** Violin plots of the volume distribution of all analyzed in vivo high-and low-[Na^+^] cysts. y-axis is displayed in log scale. The total number of high-[Na^+^] cysts (N_high_ = 1467) was 1.8 times higher than the total number of low-[Na^+^] cysts (N_low_ = 832). High-[Na^+^] cysts showed a significantly higher volume than low-[Na^+^] cysts (3.3 mL vs. 2.8 mL, p<0.05, Mann-Whitney U test). The largest low-[Na^+^] cyst had a volume of 72 mL. Above this threshold only high-[Na^+^] cysts were detected in our cohort. **B** Percentage of low-[Na^+^] cysts depending on the location of the cysts within the kidneys. Low-[Na^+^] cysts are indicated in blue, and high-[Na^+^] cysts in red. The percentage of low-[Na^+^] cysts was assessed for the whole kidney as well as for the inner and outer regions of the kidney, each defined as 50% of total kidney volume. Based on the center of the cyst mask, cysts were allocated to the inner or the outer part of the kidney. No significant difference of the percentage of low-[Na^+^] cysts in the two evaluated (center/out) regions could be found (p > 0.05, Mann-Whitney U test).

To determine whether high-[Na^+^] cysts are mainly located in the renal cortex and low-[Na^+^] cysts primarily in the renal medulla, as suggested by their origin, we assigned the cysts to their renal anatomical site. In advanced ADPKD, the anatomical compartments of the kidney cannot be sufficiently distinguished due to the cystic transformation. We therefore defined an outer part and an inner part of the kidney based on half of the kidney volume and analyzed the number of the two cyst types in each compartment. No specific localization pattern of the two cyst entities could be found (n_low,total_ = 40% (IQR 11%-54%), n_low,out_ = 35% (IQR 10%-59%), n_low,center_ = 41% (IQR 9%-56%))(**Fig. 4B**).

## DISCUSSION

Cysts in ADPKD represent a heterogeneous population but current clinical imaging technology solely focuses on kidney volume (*12*), as it is neither able to determine cyst composition nor its origin. The key achievement of our study is the first non-invasive, in vivo classification of cysts in ADPKD patients based on their [Na^+^] using 7 Tesla ^23^Na MRI. Furthermore, we could validate our measurements by comparing ^23^Na MRI results with direct examination of cyst fluid from nephrectomized ADPKD kidneys. Histological analysis of extracted cysts from these kidneys showed a clear association between cyst [Na^+^] and tubular origin with high-[Na^+^] linked to proximal tubule and low-[Na^+^] to distal tubule and collecting duct.

Assessment of nephrectomized ADPKD kidneys by ^23^Na MRI and subsequently by direct determination of electrolytes in cyst fluid demonstrated two distinct populations of cysts, either with [Na^+^] in the range of serum or with very low-[Na^+^]. This finding is consistent with early observations from the 1950s, when Bricker and Patton first reported markedly low Na^+^ concentrations in ADPKD cyst fluid (*13*). Subsequent work in the 1960s and 1970s, particularly by Gardner and colleagues, provided a more detailed characterization of cyst electrolyte composition and proposed that cyst [Na⁺] reflects proximal or distal tubular function (*7*). Later studies further supported this concept by demonstrating distinct electrical properties of low-and high-[Na⁺] cysts, consistent with tight versus leaky epithelia (*9*). In addition, low-and high-[Na^+^] cysts were categorized as distal or proximal based on physiological considerations (*6*). Despite extensive research in ADPKD, to our knowledge no prior study has assessed cyst epithelial markers in relation to cyst [Na^+^]. Not all cyst epithelia examined showed staining for specific tubule markers, which is a phenomenon also found by other investigators (*14*). In a prospective analysis of cyst origin in a mouse model of PKD1, the proportion of cysts lacking labelling for tubular markers increased from 2% to 15% after one year, indicating dedifferentiation of cysts with disease progression (*15*). Corresponding to these observations staining failure occurred mainly in high-[Na^+^] cysts known to be leaky and which might increase in number with age as implied by our in vivo measurements. The mechanism underlying the age-dependent increase in high-[Na⁺] cysts remains unclear, but may relate to cumulative epithelial alterations, intermittent inflammatory events, or repeated microinjuries over time. Importantly, none of the cysts in our cohort showed double staining of proximal and distal tubule markers indicating a clear attribution to a tubule segment. This observation is in line with a study of human ADPKD cysts, in which a mutually exclusive expression of aquaporin-1 (a marker for proximal tubule) and aquaporin-2 (a marker for collecting duct) in cysts of patients with early as well as end-stage ADPKD was demonstrated (*16*). Since we could solely find vasopressin V2 receptors in low-[Na^+^] cysts, the vasopressin V2 receptor antagonist tolvaptan - the only approved therapeutic agent in ADPKD - is supposed to inhibit the secretion and growth of this cyst type exclusively. While this mechanism is physiologically well supported by the segment-specific expression of V2R, its clinical relevance for cyst-type–specific treatment effects has not yet been prospectively validated.

Contrary to prevailing assumptions that ∼70% of cysts derive from the collecting duct (*14*), our in vivo data demonstrate substantial interindividual heterogeneity in cyst origin and [Na⁺] distribution. On the one hand we found patients with up to 91.6% of high-[Na^+^] cysts, on the other hand, in some patients, low-[Na^+^] cysts predominated, with a maximum proportion of 83.9%. When considering all cysts examined, high-[Na^+^] cysts were significantly larger than low-[Na^+^] cysts and the number of high-[Na^+^] cysts exceeded low-[Na^+^] cysts by 1.8-fold. This observation is remarkable because - at least in our cohort - it suggests that the majority of cysts originate from the proximal tubule and according to our ex vivo tubule staining do not express vasopressin V2 receptors. Corresponding to our observations, former puncture studies found a preponderance of high-[Na^+^] cysts and postulated a high degree of individual heterogeneity in distribution of the two cyst types (*8*).

In our in vivo study, the cyst type could not be predicted by cyst localization or cyst volume and therefore could not be extrapolated from the morphological ^1^H MRI assessment. Our ^23^Na MRI setup was able to directly visualize the cyst Na^+^ composition in ADPKD kidneys thereby adding functional information to the hitherto solely morphological images. The lack of a distinct cortical or medullary localization pattern does not contradict the functional assignment of high-and low-[Na⁺] cysts to proximal and distal/collecting duct origin. In advanced ADPKD, the profound distortion of renal architecture disrupts the native corticomedullary organization, and cyst expansion, displacement, and coalescence obscure the anatomical position of the originating nephron segment. As a result, the final spatial location of a cyst within the kidney no longer reflects its tubular origin. This explains why cyst type could not be inferred from anatomical position or cyst volume on conventional ¹H MRI, underscoring the added value of ²³Na MRI in providing functional information beyond morphology. ^23^Na MRI has been applied before to assess the corticomedullary Na^+^ gradient of the kidney in humans (*17–21*). Furthermore, Lemoine et al. performed in vivo ^23^Na MRI measurements of kidneys from four ADPKD patients reporting a wide range of kidney cystic Na^+^ content (*22*). They used a 3 Tesla MRI system, which required longer acquisition times and larger voxel volumes. In contrast, the implementation of ^23^Na MRI at 7 Tesla enabled shorter acquisition times and a higher spatial resolution. Combined with advanced post-processing techniques, this enabled classification into low-and high-[Na^+^] cysts and quantitative analysis of cyst-type distribution on an individual basis.

Tolvaptan is able to slow cyst growth and eGFR-decline and thereby disease progression, however individual response rates of ADPKD patients to tolvaptan treatment in the approval studies are unclear (*10, 23, 24*). In a study evaluating the efficacy of tolvaptan using changes in TKV growth rate during treatment, 44% of patients showed a reduction in annual TKV growth of less than 1% and were therefore classified as “non-responders” (*25*). This indicates that a substantial proportion of ADPKD patients will derive little or no benefit from tolvaptan therapy. Currently, however, no methods are available to predict tolvaptan efficacy at the individual patient level. If the hypothesis is confirmed that tolvaptan primarily inhibits the growth of low-[Na^+^] cysts, ^23^Na MRI could potentially serve as a tool to predict treatment response in individual patients. Furthermore, as new therapeutic approaches for ADPKD are being developed, tools capable of assessing their therapeutic effects on the two cyst types would be highly valuable (*26*).

We are aware of several limitations of our study. First, the relatively low number of participants in our clinical study limits statistical power for subgroup analyses, such as tolvaptan treatment, genetic background and gender aspects. Second, our pilot study is susceptible to selection bias and unmeasured confounding factors. Thus, our results must be confirmed in a larger ADPKD patient cohort. Third, due to the limited spatial resolution of in vivo ^23^Na MRI (6 mm isotropic), only cysts with a volume greater than 1 mL were evaluated, which might introduce a bias in the detected ratio of low-[Na^+^] vs. high-[Na^+^] cysts. Obtaining reliable information for smaller cysts remains challenging, as increasing spatial resolution would substantially prolong scan times. However, in future, improved radiofrequency coil setups and iterative post-processing methods based on the morphological MR images might overcome this limitation. Fourth, cysts that are outside the boundaries of the B_1_ calibration phantom were excluded from analysis, which might introduce an exclusion bias in the assessment of very large cystic kidneys. However, this affects less than 4% of overall assessed cysts. Fifth, in our in vivo measurements we solely performed a classification into low-[Na^+^] and high-[Na^+^] based on a threshold. Determination of absolute [Na^+^] might reveal additional insights. With the used setup, calibration of the ^23^Na signal by external references was not possible due to the limited space within the coil. However, according to our ex vivo data of nephrectomized ADPKD kidneys and previous puncture studies a clear threshold between low-[Na^+^] and high-[Na^+^] cysts exists (*11*). As cyst [Na^+^] is no continuous variable, a binary classification should be sufficient to characterize cysts and their origin in ADPKD patients.

In conclusion, this study is the first to characterize and categorize cysts into low-and high-[Na⁺] in ADPKD patients in vivo using ^23^Na MRI. The method was validated by ex vivo cyst fluid analysis enabling assignment to cyst origin based on [Na^+^]. We observed substantial interindividual heterogeneity in cyst [Na⁺] distribution, independent of total kidney volume, annual growth rate or anatomical localization. This heterogeneity may carry prognostic relevance and suggests that future therapies should consider the individual cyst profile. Tolvaptan may primarily target vasopressin V2 receptor-positive low-[Na^+^] cysts and emerging compounds may require tailored application depending on the underlying tubular origin of cysts. Our work establishes functional cyst profiling by ^23^Na MRI as a new dimension in ADPKD imaging, with potential implications for patient stratification and personalized therapy.

## MATERIAL AND METHODS

### Clinical study

We performed an ex vivo and an in vivo study. In the ex vivo study, eight kidneys from ADPKD patients who underwent nephrectomy either due to recurrent cyst infection or due to suspected renal carcinoma were assessed by ^23^Na MRI. Specific cysts with high or low-[Na^+^], as identified by ^23^Na MRI, were punctured, and electrolyte concentrations, osmolality and further metabolic parameters from cyst fluid were analyzed in our clinical laboratory.

To classify cyst [Na^+^] in vivo, an observational study was conducted that included 21 ADPKD patients. One patient was excluded because kidney size precluded adequate ^23^Na MRI assessment (TKV = 11.0 L, htTKV = 5830 mL/m). Sixty minutes before MRI measurements, patients received a water load (15 mL/kg body weight) to attenuate the renal cortico-medullary Na^+^ gradient, thereby reducing possible interference with cyst [Na^+^] classification (*18*). Additionally, blood pressure was measured and blood as well as urine samples were obtained. Inclusion criteria for patients were age ≥ 18 years and clinical or genetic diagnosis of ADPKD. Patients with pacemakers, metal implants or other MRI contraindications were excluded. We focused on patients with remaining kidney function and thus excluded patients on dialysis or after kidney transplantation. All MRI scans were conducted between August 2024 and April 2025 at the Institute of Radiology, University Hospital of Erlangen. The local Ethics Committees of the University of Erlangen-Nürnberg, Germany (clinical trial: No. 22-271-Bm) and the Ethics Committees of University of Regensburg, Germany (assessment of nephrectomized kidneys: No. 20-1886_7-101) approved the studies, which were conducted according to the declaration of Helsinki principles. All participants provided written informed consent.

### Measurement setup

The ^23^Na MRI measurements were performed using a 7 Tesla whole-body MRI system (MAGNETOM Terra.X, Siemens Healthcare, Erlangen, Germany). ^23^Na MR images of the nephrectomized kidneys were acquired with a double-resonant ^23^Na/^39^K birdcage calf coil (Rapid Biomedical, Rimpar, Germany) (*27*). A reference holder with known Na^+^ concentrations (*c_ref_* = [30, 60, 90, 120, 150] mM) was incorporated into the setup to retrospectively calibrate the ^23^Na signal of the ex vivo kidney cysts. For anatomical ^1^H MR images of the ex vivo kidneys, a ^1^H/^31^P RF birdcage coil (Rapid Biomedical, Rimpar, Germany) was used. The shared coil holder for both ²³Na and ¹H acquisitions allowed the nephrectomized kidney to remain in the same position throughout the entire examination. The coil setup for the in vivo patient measurements included a ^23^Na birdcage coil that was combined with two 4Tx/8Rx ^1^H torso arrays, allowing for interleaved ^23^Na/^1^H imaging, which included parallel transmission (pTx) for improved FA homogeneity for ^1^H, as described by Ruck et al. (*28*).

All patients underwent an additional ^1^H MRI scan at 3 Tesla (MAGNETOM VIDA, Siemens Healthineers, Erlangen, Germany) on the same day to obtain high-resolution anatomical images with good cyst contrast for volumetric analysis and individual cyst segmentations.

### Ex vivo ^23^Na MRI protocol

The ^23^Na images were acquired with a density-adapted 3D radial sequence (*29*) ((*Δ*x_23Na_)^3^ = (3 mm)^3^, FA_23Na_ = 82°, TR_23Na_ = 60 ms, TE_23Na_ = 0.4 ms, TA = 20 min). To correct flip angle (FA) inaccuracies arising from the coil profile, an additional ^23^Na FA map was acquired using an actual flip angle imaging (AFI) sequence (*30*). After a coil change, an^1^H T_1_ weighted FLASH image was acquired ((*Δ*x_1H_)^3^ = (1×1×3 mm)^3^, FA_1H_ = 40°, TR_1H_ = 8.1 ms, TE_1H_ = 3.47 ms, TA = 3:43 min) and linear interpolated to 1 mm in slice direction. Owing to its higher in plane resolution, this image was used for cysts segmentation. After puncturing selected cysts under ^23^Na MRI guidance, a diluted contrast agent was injected into these cysts to locate them in a second T1 weighted FLASH image. The sequence parameters are listed in Supplementary Table 1 with the note “ex vivo”.

### In vivo ^23^Na MRI protocol

ADPKD patients underwent two MRI scans. Interleaved **^1^**H(pTx)/^23^Na MRI was performed at the aforementioned 7 Tesla MRI system (nominal resolution: (*Δ*x_23Na_)^3^ = (6 mm)^3^, (*Δ*x_1H_)^3^ = (2 mm)^3^, FA_23Na_ = 82°, FA_1H_ = 10°, TR_23Na_ = 60 ms, TE_23Na_ = 1.15 ms, TA = 15 min) (*28*). The measurements at 7 Tesla were conducted using a 3D density-adapted radial acquisition scheme (*29*), which is robust to motion and allows for free breathing during the scan (*31*). For homogeneous FA distribution of ^1^H imaging a universal kidney phase shim was applied (*32*). An additional AFI scan was performed to correct for transmit field inhomogeneities. The data were reconstructed offline using a custom reconstruction pipeline with additional correction for gradient non-linearity effects (*33*). In the second scan, anatomical images for segmentations were acquired at a 3 Tesla scanner (MAGNETOM Vida, Siemens Healthcare, Erlangen, Germany) applying a T_2_-weighted Half-Fourier Acquisition Single-shot Turbo Spin Echo (HASTE) sequence ((*Δ*x)^3^ = 1×1×4 mm^3^, FA = 120°, TE = 88 ms, TR = 970 ms, TA = 53 s, breath hold, with fat saturation). The sequence parameters of the in vivo measurements from 7 Tesla and 3 Tesla are also listed in Supplementary Table 1.

### MRI Data postprocessing

#### Reconstruction and B_1_ correction

Ex vivo: The Na^+^ images were reconstructed offline using a custom MATLAB script, applying a Hamming filter and a nonuniform fast Fourier transform as well as an interpolation to 1 mm isotropic voxel size (*34, 35*). Additionally, a pixelwise FA correction was applied using the FA map obtained by the AFI measurement. ^1^H images were rigidly registered to the ^23^Na images using Elastix to compensate for a potential misalignment of both images after the coil change (*36*). Manual segmentations were performed on the high-resolution ^1^H image, including the reference holders and the punctuated cysts using the VolumeSegmenter App in MATLAB.

In vivo: The ^23^Na and ^1^H images of the ADPKD patients acquired at 7 Tesla were reconstructed offline using the same custom-written reconstruction script as above, as well as a Hamming filter. ^23^Na and ^1^H images were interpolated to an isotropic voxel size of 1 mm. The anatomical T_2_-weighted HASTE ¹H images served as the basis for the segmentations. The FA profile of Na^+^ images was corrected pixelwise with the subject specific FA map obtained from the AFI measurement. Additionally, inhomogeneities from the receive profile were corrected with a B_1_^-^-map acquired from a phantom measurement, mimicking the dielectric properties from tissue (*28*). The correction for transmit and receive field of the coil was calculated pixelwise as follows (*37*):

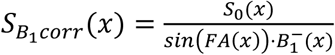

The in vivo ^1^H images from 7 Tesla and from 3 Tesla were registered in a two-iteration approach using Elastix applying a rigid transformation. In the first step the entire image was registered. The parameters obtained from the first iteration were used for the second iteration as initial values. In this second step, individual masks of each kidney were used to perform separate registration for the left and right kidney. Individual cysts were segmented on the ^1^H T_2_-weighted HASTE images using the volumeSegmenter in MATLAB. Cysts that were not completely covered by the phantom-based B_1_^-^ map were excluded, as a B_1_^-^ correction in these cysts was not possible.

### Partial volume correction and classification into high-and low-[Na^+^] in vivo

In ^23^Na MRI partial volume effects (PVE) impair signal evaluation due to the low spatial resolution and blurring from the point spread function (PSF). To address this limitation, a partial volume correction (PVC) was applied following an approach based on the geometric transfer matrix (GTM) approach as presented by Niesporek et al. (*38*). To estimate the partial volume effect (PVE) from the surrounding tissue, each cyst segmentation was dilated by 10 mm in all spatial directions. This radius corresponds to the 2σ of the PSF, characterizing the effective spatial blurring of the ^23^Na acquisition. The mean signal intensity within the resulting shell region was subsequently calculated. For each cyst and its environment, a partial volume correction was performed. Due to the PVE, signal from high-[Na^+^] cysts spreads into adjacent tissue, which mostly has a lower [Na^+^] than these cysts. Thus, the PVC results in an upward correction of the mean ^23^Na signal in these cysts. In contrast, for low-[Na^+^] cysts, the surrounding tissue typically shows a higher [Na^+^] and therefore the PVE increases the ^23^Na signal in these cysts. Accordingly, application of the PVC results in a downward correction of the mean ^23^Na signal of such cysts. Therefore, the difference of the mean ^23^Na signal of each cyst before and after PVC was used to distinguish between high and low-[Na^+^] cysts.

### Localization of low-[Na^+^] cysts

In a further step, the spatial distribution of the low-[Na^+^] cysts in the kidneys was evaluated. The total kidney mask was isotropically reduced in size using a uniform 3D morphological erosion and subdivided into an inner and an outer region, such that the volume ratio between the outer and inner region was 50 ± 2%. Each cyst was assigned to either the inner or the outer region according to the center of gravity of its segmentation mask. The ratio of low-[Na^+^] cysts was computed based on the number of cysts in each region.

### Direct assessment of cyst fluid

Electrolytes, albumin, glucose, uric acid, urea and creatinine of aspired cyst fluid were directly assessed by our clinical laboratory using a chemistry analyzer (Beckman Coulter DxC 700 AU and AU5800 Chemistry Analyzer, Brea, USA). Alpha-1-microglobulin was determined by nephelometry (Atellica NEPH 630, Siemens, Germany) and osmolality was measured by freezing point osmometry (OsmoPRO, Advanced Instruments, USA).

### Histochemistry of cyst epithelia

Assessed cysts were dissected and stained for markers of the proximal tubule, distal tubule and collecting duct. Kidney tissue from adult nephrectomy specimens was fixed with 4% paraformaldehyde. Paraffin sections of 2.5 μm thickness were deparaffinized in xylene and rehydrated through graded ethanol. Endogenous peroxidase activity was blocked with 3% H_2_O_2_. To retrieve antigens, sections were boiled in a Biocare Decloaking Chamber using Target Retrieval Solution (Dako, Glostrup, Denmark) at pH 6 for AQP2 and NCC and pH 9 for SGLT2 and V2R. The sections were blocked with an Avidin/Biotin Blocking Kit (Vector Laboratories, CA, USA) and washed in Tris-buffered saline (TBS, pH 7.4). Primary antibodies (Supplementary Table 2) were applied overnight at 4°C in TBS with 1% bovine serum albumin (BSA). After washing, sections were incubated with a secondary antibody for one hour at room temperature in TBS with 1% BSA (anti-rabbit biotinylated IgG, BA-1000, Vector Laboratories, Burlingame, CA; 1:500). Signals were amplified by the use of the Vectastain Elite ABC Kit (Vector Laboratories, Burlingame, CA) according to the manufacturer’s instructions. Detection was performed with ready-to-use streptavidin-HRP (Abcam, UK) and signals were developed with DAB (3,3’ – diaminobenzidine; Dako, Glostrup, Denmark). Sections were counterstained with haematoxylin (Carl Roth, Karlsruhe, Germany). Coverslips were mounted with a hydrophilic mounting agent (Aquatex, Sigma-Aldrich, MO, USA). Photos were acquired using a DM6000B brightfield microscope (Leica, Wetzlar, Germany) and a Leica DFC 450C camera.

### Clinical assessment

Blood pressure was measured after 5 min of rest in seated position using an automated oscillometric device (Dinamap, Critikon, Carlsbad, USA) prior to MRI assessment. Blood pressure was analyzed on both upper arms and the arm with the higher pressure was chosen. Three consecutive measurements were averaged. Prior to MRI measurements, venous blood and urine samples were obtained and stored at-80°C.

Mayo classification was determined using the htTKV, which was obtained by segmentation using the nnU-Net integration in MITK nnInteractive (*39*).

## Statistics

All data were analyzed by IBM SPSS Statistics (Version 31). The Shapiro-Wilk-test was used to assess the distribution of our data. All normally distributed data were subsequently analyzed using the independent Student’s t-test or paired t-test, data with a skewed distribution were analyzed using the Mann-Whitney U test (independent samples) or Wilcoxon signed-rank test (paired samples). Results were expressed as mean ± standard deviation for normally distributed data and as median and interquartile range for the data lacking a normal distribution. Spearman rank correlation was used to compute the correlation coefficient. A p-value < 0.05 was considered significant. Two-sided tests of hypotheses were used throughout.

## List of Supplementary Materials

Supplementary Table 1: Sequence parameters for 7 Tesla and 3 Tesla MRI measurements

Supplementary Table 2: Primary antibodies used for staining of tubule markers

Supplementary Table 3: In vivo cyst evaluation of each patient, complete data set

Supplementary Figure 1: Additional examples of low-and high-[Na^+^] cysts with staining of tubular origin markers

## Data Availability

All data produced in the present study are available upon reasonable request to the authors.

## Acknowledgments

We sincerely thank our study participants and our study nurses Manuela Linß, Michaela Arend and Lina Voß.

## Author contributions

Conceptualization: JS, AD, BB, AMN, CK

Methodology: JS, LR, KT, PL, JMH, RU, AK, AMN

Investigation: JS, AD, PL, KT, JMH, SH, BN, MS, AK, RU, CK

Visualization: JS, BB

Funding acquisition: AD, SH, MS, BB, CK, AMN

Project administration: BB, AMN, CK

Supervision: MU, MS, BW, BB, AMN, CK

Writing – original draft: JS, CK

Writing – review & editing: all authors

## Data and materials availability

All data are available upon request.

## References

1. V. E. Torres, P. C. Harris, Y. Pirson, Autosomal dominant polycystic kidney disease. Lancet 369, 1287–1301 (2007).

2. F. T. Chebib, C. Hanna, P. C. Harris, V. E. Torres, N. K. Dahl, Autosomal Dominant Polycystic Kidney Disease: A Review. JAMA 333, 1708–1719 (2025).

3. J. J. Grantham, J. L. Geiser, A. P. Evan, Cyst formation and growth in autosomal dominant polycystic kidney disease. Kidney Int 31, 1145–1152 (1987).

4. Q. Li, Y. Wang, W. Deng, Y. Liu, J. Geng, Z. Yan, F. Li, B. Chen, Z. Li, R. Xia, W. Zeng, R. Liu, J. Xu, F. Xiong, C. L. Wu, Y. Miao, Heterogeneity of cell composition and origin identified by single-cell transcriptomics in renal cysts of patients with autosomal dominant polycystic kidney disease. Theranostics 11, 10064–10073 (2021).

5. Y. Muto, E. E. Dixon, Y. Yoshimura, H. Wu, K. Omachi, N. Ledru, P. C. Wilson, A. J. King, N. Eric Olson, M. G. Gunawan, J. J. Kuo, J. H. Cox, J. H. Miner, S. L. Seliger, O. M. Woodward, P. A. Welling, T. J. Watnick, B. D. Humphreys, Defining cellular complexity in human autosomal dominant polycystic kidney disease by multimodal single cell analysis. Nat Commun 13, 6497 (2022).

6. R. Huseman, A. Grady, D. Welling, J. Grantham, Macropuncture study of polycystic disease in adult human kidneys. Kidney Int 18, 375–385 (1980).

7. K. D. Gardner, Jr., Composition of fluid in twelve cysts of a polycystic kidney. N Engl J Med 281, 985–988 (1969).

8. K. D. Gardner, Jr., J. S. Burnside, B. J. Skipper, S. K. Swan, W. M. Bennett, B. A. Connors, A. P. Evan, On the probability that kidneys are different in autosomal dominant polycystic disease. Kidney Int 42, 1199–1206 (1992).

9. R. D. Perrone, In vitro function of cyst epithelium from human polycystic kidney. J Clin Invest 76, 1688–1691 (1985).

10. V. E. Torres, A. B. Chapman, O. Devuyst, R. T. Gansevoort, J. J. Grantham, E. Higashihara, R. D. Perrone, H. B. Krasa, J. Ouyang, F. S. Czerwiec, T. T. Investigators, Tolvaptan in patients with autosomal dominant polycystic kidney disease. N Engl J Med 367, 2407– 2418 (2012).

11. S. Terryn, A. Ho, R. Beauwens, O. Devuyst, Fluid transport and cystogenesis in autosomal dominant polycystic kidney disease. Biochim Biophys Acta 1812, 1314–1321 (2011).

12. M. V. Irazabal, L. J. Rangel, E. J. Bergstralh, S. L. Osborn, A. J. Harmon, J. L. Sundsbak, K. T. Bae, A. B. Chapman, J. J. Grantham, M. Mrug, M. C. Hogan, Z. M. El-Zoghby, P. C. Harris, B. J. Erickson, B. F. King, V. E. Torres, C. Investigators, Imaging classification of autosomal dominant polycystic kidney disease: a simple model for selecting patients for clinical trials. J Am Soc Nephrol 26, 160–172 (2015).

13. N. S. Bricker, J. F. Patton, Cystic disease of the kidneys; a study of dynamics and chemical composition of cyst fluid. Am J Med 18, 207–219 (1955).

14. R. R. Verani, F. G. Silva, Histogenesis of the renal cysts in adult (autosomal dominant) polycystic kidney disease: a histochemical study. Mod Pathol 1, 457–463 (1988).

15. K. Hopp, C. J. Ward, C. J. Hommerding, S. H. Nasr, H. F. Tuan, V. G. Gainullin, S. Rossetti, V. E. Torres, P. C. Harris, Functional polycystin-1 dosage governs autosomal dominant polycystic kidney disease severity. J Clin Invest 122, 4257–4273 (2012).

16. O. Devuyst, C. R. Burrow, B. L. Smith, P. Agre, M. A. Knepper, P. D. Wilson, Expression of aquaporins-1 and-2 during nephrogenesis and in autosomal dominant polycystic kidney disease. Am J Physiol 271, F169–183 (1996).

17. J. T. Grist, F. Riemer, E. S. S. Hansen, R. S. Tougaard, M. A. McLean, J. Kaggie, N. Bogh, M. J. Graves, F. A. Gallagher, C. Laustsen, Visualization of sodium dynamics in the kidney by magnetic resonance imaging in a multi-site study. Kidney Int 98, 1174–1178 (2020).

18. A. Akbari, S. Lemoine, F. Salerno, T. L. Marcus, T. Duffy, T. J. Scholl, G. Filler, A. A. House, C. W. McIntyre, Functional Sodium MRI Helps to Measure Corticomedullary Sodium Content in Normal and Diseased Human Kidneys. Radiology 303, 384–389 (2022).

19. S. Haneder, S. Konstandin, J. N. Morelli, A. M. Nagel, F. G. Zoellner, L. R. Schad, S. O. Schoenberg, H. J. Michaely, Quantitative and qualitative (23)Na MR imaging of the human kidneys at 3 T: before and after a water load. Radiology 260, 857–865 (2011).

20. N. Maril, Y. Rosen, G. H. Reynolds, A. Ivanishev, L. Ngo, R. E. Lenkinski, Sodium MRI of the human kidney at 3 Tesla. Magn Reson Med 56, 1229–1234 (2006).

21. G. Chen, Z. Liao, S. Ma, P. Luo, B. Deng, X. Zhang, Q. Wang, H. Tang, X. Lu, X. Hu, N. Gong, Z. Li, Functional Sodium MRI in the Measurement of Corticomedullary Sodium Content and Its Role in Differentiating Between Transplanted Kidneys With Superior and Inferior Graft Function. J Magn Reson Imaging 62, 1209–1216 (2025).

22. S. Lemoine, A. Akbari, C. W. McIntyre, (23)NaMRI Assessed Cyst Sodium Concentration in Polycystic Kidney Disease to Identify Cyst Metabolic Activity: A Proof of Concept Study. Kidney Med 6, 100820 (2024).

23. V. E. Torres, A. B. Chapman, O. Devuyst, R. T. Gansevoort, R. D. Perrone, G. Koch, J. Ouyang, R. D. McQuade, J. D. Blais, F. S. Czerwiec, O. Sergeyeva, R. T. Investigators, Tolvaptan in Later-Stage Autosomal Dominant Polycystic Kidney Disease. N Engl J Med 377, 1930–1942 (2017).

24. V. E. Torres, A. B. Chapman, O. Devuyst, R. T. Gansevoort, R. D. Perrone, A. Dandurand, J. Ouyang, F. S. Czerwiec, J. D. Blais, T. T. Investigators, Multicenter, open-label, extension trial to evaluate the long-term efficacy and safety of early versus delayed treatment with tolvaptan in autosomal dominant polycystic kidney disease: the TEMPO 4:4 Trial. Nephrol Dial Transplant 33, 477–489 (2018).

25. H. Dev, Z. Hu, J. D. Blumenfeld, A. Sharbatdaran, Y. Kim, C. Zhu, D. Shimonov, J. M. Chevalier, S. Donahue, A. Wu, A. RoyChoudhury, X. He, M. R. Prince, The Role of Baseline Total Kidney Volume Growth Rate in Predicting Tolvaptan Efficacy for ADPKD Patients: A Feasibility Study. J Clin Med 14, (2025).

26. A. S. L. Yu, R. Garg, K. A. Bellovich, A. L. Silva, F. T. Chebib, C. Padgett, E. C. Y. Lee, T. M. Valencia, T. L. Kline, A. Gregory, K. Carroll, V. Patel, Farabursen Increases Urinary Polycystin-1 and Polycystin-2 and Reduces Height-Adjusted Total Kidney Volume Growth in Patients with ADPKD: SA-OR089. Journal of the American Society of Nephrology 36, 10.1681/ASN.202527ntvtc202523 (2025).

27. L. V. Gast, S. Volker, M. Utzschneider, P. Linz, T. Wilferth, M. Muller, C. Kopp, B. Hensel, M. Uder, A. M. Nagel, Combined imaging of potassium and sodium in human skeletal muscle tissue at 7 T. Magn Reson Med 85, 239–253 (2021).

28. L. Ruck, N. Egger, T. Wilferth, J. Schirmer, L. V. Gast, S. Nagelstrasser, S. Wildenberg, A. Bitz, T. Lanz, T. Platt, S. Konstandin, C. Kopp, M. Uder, A. M. Nagel, Interleaved 23Na/1H MRI of the human heart at 7 T using a combined 23Na/1H coil setup and 1H parallel transmission. Magnet Reson Med 94, 231–241 (2025).

29. A. M. Nagel, F. B. Laun, M. A. Weber, C. Matthies, W. Semmler, L. R. Schad, Sodium MRI using a density-adapted 3D radial acquisition technique. Magn Reson Med 62, 1565– 1573 (2009).

30. V. L. Yarnykh, Actual flip-angle imaging in the pulsed steady state: a method for rapid three-dimensional mapping of the transmitted radiofrequency field. Magn Reson Med 57, 192–200 (2007).

31. G. H. Glover, J. M. Pauly, Projection Reconstruction Techniques for Reduction of Motion Effects in Mri. Magnet Reson Med 28, 275–289 (1992).

32. N. Egger, S. Nagelstrasser, S. Wildenberg, A. Bitz, L. Ruck, J. Herrler, C. R. Meixner, R. Kimmlingen, T. Lanz, S. Schmitter, M. Uder, A. M. Nagel, Accelerated B1+ mapping and robust parallel transmit pulse design for heart and prostate imaging at 7 T. Magn Reson Med 92, 1933–1951 (2024).

33. S. J. Doran, L. Charles-Edwards, S. A. Reinsberg, M. O. Leach, A complete distortion correction for MR images: I. Gradient warp correction. Phys Med Biol 50, 1343–1361 (2005).

34. I. The MathWorks. (The MathWorks, Inc., Natick, MA, USA, 2024), vol. R2024b.

35. J. A. Fessler, B. P. Sutton, Nonuniform fast Fourier transforms using min-max interpolation. Ieee T Signal Proces 51, 560–574 (2003).

36. S. Klein, M. Staring, K. Murphy, M. A. Viergever, J. P. Pluim, elastix: a toolbox for intensity-based medical image registration. IEEE Trans Med Imaging 29, 196–205 (2010).

37. L. Ruck, N. Egger, B. Zobler, J. Schirmer, S. Nagelstrasser, A. Bitz, T. Platt, S. Konstandin, C. Kopp, M. Uder, A. M. Nagel, Improved Myocardial Sodium Quantification at 7 T Using Interleaved (23)Na/(1)H pTx MRI With Motion and Anatomy-Based B(1) Correction. Magn Reson Med 96, 173–190 (2026).

38. S. C. Niesporek, S. H. Hoffmann, M. C. Berger, N. Benkhedah, A. Kujawa, P. Bachert, A. M. Nagel, Partial volume correction for in vivo (23)Na-MRI data of the human brain. Neuroimage 112, 353–363 (2015).

39. F. Isensee, Rokuss, M.*, Krämer, L.*, Dinkelacker, S., Ravindran, A., Stritzke, F., Hamm, B., Wald, T., Langenberg, M., Ulrich, C., Deissler, J., Floca, R., & Maier-Hein, K., nnInteractive: Redefining 3D Promptable Segmentation. https://arxiv.org/abs/2503.08373, (2025).

