## Supplementary Material for "Non-invasive ²³Na MRI Identifies Functionally Distinct Cyst Phenotypes in Autosomal Dominant Polycystic Kidney Disease"

**Supplementary Table 1: Sequence parameters for 7 Tesla and 3 Tesla MRI measurements**

| | 7 Tesla | | | | | | 3 Tesla |
|---|---|---|---|---|---|---|---|
| | Interleaved $^{23}$Na/$^{1}$H | | $^{23}$Na | $^{23}$Na AFI | | $^{1}$H GRE | $T_2$-weighted HASTE |
| **Application** | in vivo | | ex vivo | in vivo | ex vivo | ex vivo | in vivo |
| **Nucleus** | $^{23}$Na | $^{1}$H | $^{23}$Na | $^{23}$Na | $^{23}$Na | $^{1}$H | $^{1}$H |
| **Nominal Resolution [mm]** | 6x6x6 | 2x2x2 | 3x3x3 | 12x12x12 | 6x6x6 | 1x1x3 | 1x1x4 |
| **Nominal FA [°]** | 82 | 2 | 82 | 60 | 60 | 40 | 120 |
| **Projections / Slices** | 15 000 centre-out: half proj. | 60 000 centre-through: full proj. | 20 000 centre out: full proj. | 3000 centre out: full proj. | 3000 centre out: full proj. | 88 slices | 38 - 62 slices* |
| **TR [ms]** | 60 | $TR_1$ = 13.08 $TR_2$ = 20.76 | 60 | $TR_1$ = 48 $TR_2$ = 12 | $TR_1$ = 48 $TR_2$ = 12 | 8.1 | 970 |
| **TE [ms]** | 1.15 | 2.5 | 0.4 | 1.05 | 0.4 | 3.47 | 88 |
| **Additional** | Density-adapted 3D radial readout | | | | | | Fat saturation |
| | Free breathing | | | | | | Breath hold |
| **TA [min]** | 15 | | 20 | 5 | 5 | 3:43 | 0:53 - 1:52 |

GRE, gradient echo; HASTE, half acquisition single-shot turbo spin echo; FA, flip angle; half/full proj., half/full radial projection, TR, repetition time; TE, echo time; TA, acquisition time; *number of acquired slices varied depending on kidney size.

**Supplementary Table 2: Primary antibodies used for staining of tubule markers**

| Target | Catalogue No. | Dilution | Host | Source |
|---|---|---|---|---|
| SGLT2 | 24654-1-AP | 1:200 | Rabbit | Thermo Fisher Scientific, MA, USA |
| NCC | SPC-402 | 1:500 | Rabbit | StressMarq Biosciences, BC, Canada |
| V2R | EPR24555-59 | 1:100 | Rabbit | Abcam, Cambridge, UK |
| AQP2 | PA5-22865 | 1:250 | Rabbit | Thermo Fisher Scientific, MA, USA |

SGLT2, sodium-glucose cotransporter-2; NCC, sodium-chloride cotransporter; V2R, vasopressin V2 receptor; AQP2, aquaporin-2.

**Supplementary Table 3: In vivo cyst evaluation of each patient, complete data set**

| **Patient ID** | **1** | **2** | **3** | **4** | **5** | **6** | **7** | **8** | **9** | **10** | **11** | **12** | **13** | **14** | **15** | **16** | **17** | **18** | **19** | **20** |
|---|---|---|---|---|---|---|---|---|---|---|---|---|---|---|---|---|---|---|---|---|
| TKV [L] | 1.38 | 0.73 | 5.58 | 2.31 | 3.27 | 1.42 | 1.18 | 2.26 | 1.42 | 4.24 | 1.11 | 3.68 | 0.60 | 6.58 | 0.98 | 3.01 | 1.31 | 0.40 | 0.86 | 2.19 |
| htTKV [mL/m] | 757 | 398 | 3101 | 1337 | 1892 | 793 | 635 | 1366 | 768 | 2436 | 698 | 2032 | 364 | 3978 | 597 | 1756 | 740 | 243 | 487 | 1326 |
| $TKV_{left}$ [L] | 0.65 | 0.44 | 3.12 | 1.24 | 1.89 | 0.85 | 0.58 | 1.18 | 0.73 | 2.37 | 0.58 | 1.96 | 0.24 | 3.44 | 0.66 | 1.32 | 0.67 | 0.21 | 0.57 | 1.24 |
| $TKV_{right}$ [L] | 0.73 | 0.28 | 2.46 | 1.06 | 1.38 | 0.58 | 0.60 | 1.08 | 0.68 | 1.87 | 0.52 | 1.72 | 0.36 | 3.15 | 0.32 | 1.69 | 0.64 | 0.19 | 0.30 | 0.95 |
| N | 106 | 31 | 241 | 156 | 239 | 65 | 64 | 146 | 84 | 211 | 83 | 244 | 23 | 155 | 32 | 211 | 58 | 12 | 49 | 89 |
| $N_{left}$ | 53 | 22 | 104 | 80 | 125 | 33 | 34 | 80 | 56 | 107 | 43 | 149 | 12 | 70 | 26 | 108 | 37 | 8 | 38 | 43 |
| $N_{right}$ | 53 | 9 | 137 | 76 | 114 | 32 | 30 | 66 | 28 | 104 | 40 | 95 | 11 | 85 | 6 | 103 | 21 | 4 | 11 | 46 |
| $N_{low}$ | 53 | 26 | 165 | 85 | 76 | 3 | 22 | 22 | 38 | 115 | 7 | 15 | 2 | 13 | 4 | 101 | 31 | 7 | 36 | 11 |
| $N_{high}$ | 53 | 5 | 76 | 71 | 163 | 62 | 42 | 124 | 46 | 96 | 76 | 229 | 21 | 142 | 28 | 110 | 27 | 5 | 13 | 78 |
| $n_{low}$ [%] | 50.0 | 83.9 | 68.5 | 54.5 | 31.8 | 4.6 | 4.4 | 15.1 | 45.2 | 54.5 | 8.4 | 6.2 | 8.7 | 8.4 | 12.5 | 47.9 | 53.5 | 58.3 | 73.5 | 12.4 |

TKV, total kidney volume; htTKV, height-adjusted total kidney volume; N, total number of evaluated cysts per patient; $N_{left/right}$, total number of evaluated cysts in left/right kidney per patient; $N_{low/high}$, total number of high- and low-[$Na^+$] cysts per patient; $n_{low}$, percentage of low-[$Na^+$] cysts.

**Supplementary Figure 1: Additional examples of low- and high-[Na$^+$] cysts with staining of tubular origin markers**

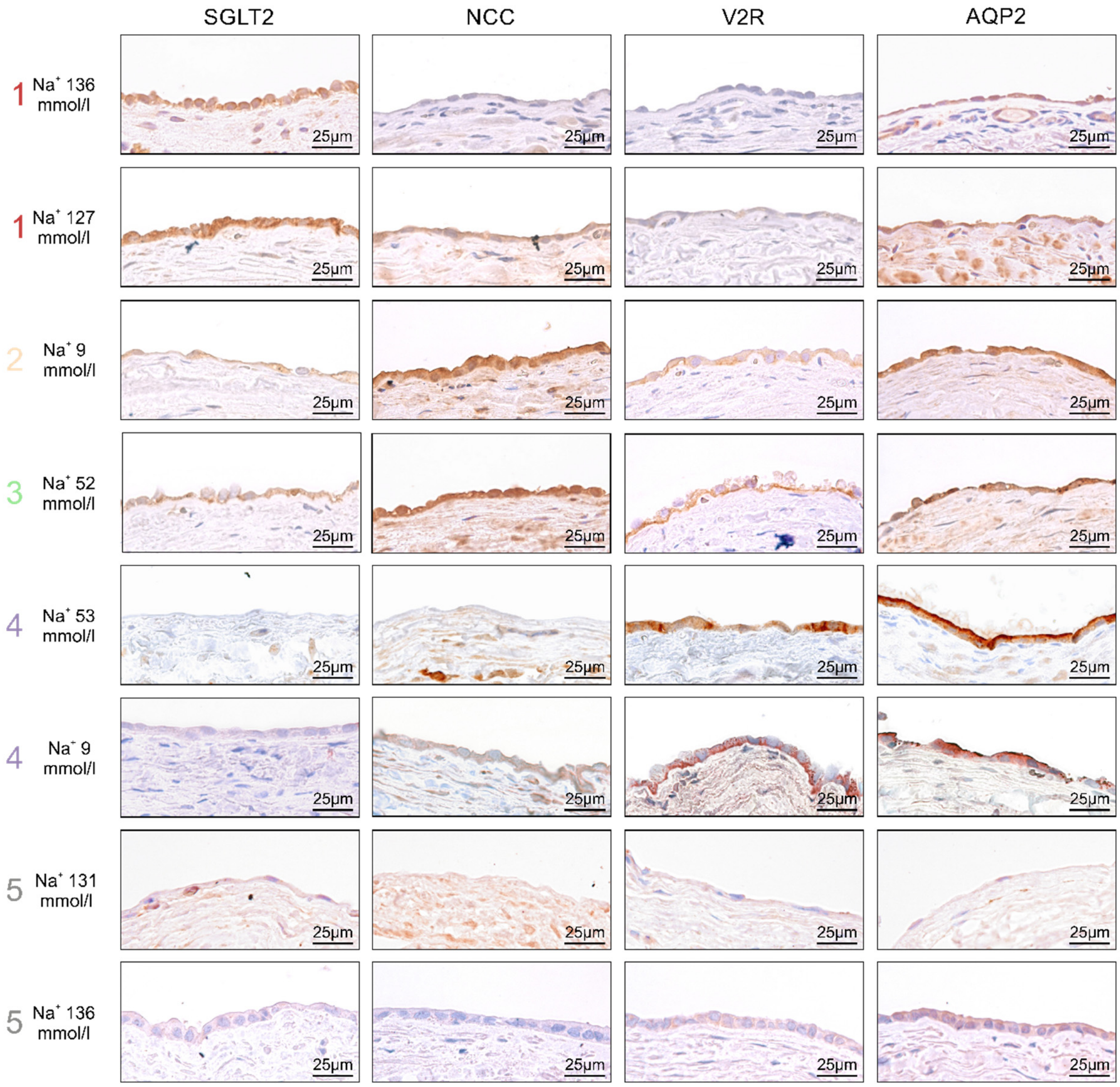


1: epithelial staining of high-[Na$^+$] cysts; 2-4: constellation of epithelial markers in low-[Na$^+$] cysts corresponding to 2: convoluted tubule, 3: connecting tubule and 4: collecting duct; 5: negative staining for tubular markers occurred predominantly in high-[Na$^+$] cysts. None of the low-[Na$^+$] cysts expressed the proximal tubular marker SGLT2. Additional proximal markers were negative in all cysts (AQP1, Megalin, Lotus lectin). No distal tubular marker could be detected in high-[Na$^+$] cysts. 40.9% of high-[Na$^+$] cysts but only 10.5% of low-[Na$^+$] cysts were negative for all tubular markers.
